# Caregiver knowledge, its determinants and its association with infant and young child feeding and water, sanitation, and hygiene practices among children with severe acute malnutrition in agrarian and pastoral settings of Ethiopia

**DOI:** 10.64898/2026.04.09.26350480

**Authors:** Mohammed Areb, Lieven Huybregts, Dessalegn Tamiru, Mariama Touré, Bayise Biru, Talla Fall, Alemayehu Haddis, Tefera Belachew

## Abstract

**Background:** This study aimed to assess caregiver knowledge of Infant and Young Child Feeding (IYCF), child health, severe acute malnutrition (SAM) screening, and Community-Based Management of Acute Malnutrition (CMAM), its determinants, and associations with IYCF/ WaSH (water, sanitation, and hygiene) practices among caregivers of children 6–59 months with SAM in Ethiopian agrarian and pastoralist settings.

**Method:** Data were from the baseline survey of the R-SWITCH Ethiopia cluster-randomized controlled trial (cRCT), which screened ∼28,000 children aged 6–59 months and identified 686 SAM cases. Caregiver knowledge was evaluated using a validated 32-item questionnaire (Cronbach’s α for internal reliability) and analyzed via linear mixed-effects and Poisson regression models in Stata 17.

**Results:** Caregiver knowledge was positively associated with improved IYCF/WaSH practices among children aged 6–23 months with SAM, including higher minimum dietary diversity (MDD: IRR=1.50), minimum acceptable diet (MAD: IRR=1.63), and reduced zero vegetable/fruit intake (IRR=0.77), as well as MDD in children aged 24–59 months, improved water access (IRR=1.19), water treatment (IRR=2.02), and handwashing stations (IRR=1.41). Literate (β = 4.1; 95% CI:1.5–6.6, p= 0.016), pregnant(β = 4.4; 95% CI:0.9–7.8, 0.018), having child weighing at a health post/ health center (β = 8.9;95% CI:3.5–14.2,p ≤ 0.001), and higher household wealth index (β = 11.8;95% CI:3.6–20.1,p= 0.005) were associated with higher knowledge, while possible depression (β = −0.3;95% CI: −0.5 to 0.0, p= 0.015) was associated with lower knowledge.

**Conclusion:** Caregiver knowledge determines better IYCF/WaSH practices among children aged 6-59 months with SAM. Literacy, pregnancy, having child weighing at a health post or health center, and greater household wealth were associated with caregivers knowledge, whereas possible depression was associated with lower knowledge. Integrating context-specific caregiver education and mental health support into CMAM, GMP(Growth monitoring and promotion), and primary care services could enhance feeding/WaSH practices in Ethiopia.

## Introduction

Malnutrition causes 35–45% of global under-5 deaths (Karlsson et al., 2022), affecting 150.2M stunted, 42.8M wasted children, including approximately 12.2M with SAM(1). In Ethiopia, 37% are stunted, 7% wasted (2). The immediate causes of malnutrition inadequate dietary intake and infection are rooted in caregiving environments marked by suboptimal IYCF, poor WaSH, and limited health access(3). Caregivers, as primary mediators of these determinants, play a decisive role. Their knowledge of nutrition and hygiene strongly predicts feeding practices; deficits in this knowledge are linked to monotonous diets, delayed introduction of nutrient-dense foods, and poor dietary habits in children under five, with maternal knowledge directly shaping children’s eating behaviors (4–7), and improvements in caregiver knowledge and practice consistently correlate with better child nutritional outcomes (8–10).

In Ethiopia, particularly in rural agrarian and pastoralist communities, gaps in caregiver knowledge remain a critical barrier to adopting WHO-recommended IYCF practices (7,11). Compounding feeding challenges, inadequate WaSH conditions also amplify malnutrition risk: the WHO estimates that 50% of undernutrition is associated with infections stemming from unsafe water, poor sanitation, or insufficient hygiene(12). Diarrheal and subclinical enteric infections common where handwashing and safe food handling are neglected suppress appetite, reduce nutrient absorption, and undermine even well-intentioned feeding efforts (13,14). Notably, caregiver health literacy and knowledge directly influences hygiene behaviors: maternal education improves handwashing at critical times (15).

This study targets children 6–59 months with SAM in Ethiopian agrarian and pastoral settings as wasting risks peak in early childhood(16). This community-based study comprehensively assessed caregiver knowledge among all 6–59-month-old children with SAM and enrolled in SAM OTPs, ensuring a population-representative sample that captures the distinct profiles of children, caregivers, and households (17). Existing research either relies on SAM cases enrolled from treatment sites which are a sample unlikely to be representative for the population(18), or fails to account for nutritional status or lacks disaggregation by treatment context an important gap(19,20). This study aimed to (1) assess caregiver knowledge across five domains (breastfeeding, complementary feeding, child health, SAM screening including MUAC/edema recognition, and CMAM) and its determinants, and (2) examine associations between caregivers knowledge and IYCF/WaSH practices, among caregivers of children aged 6–59 months screened with SAM in agrarian and pastoralist settings of Ethiopia.

## Methods

### Study setting

Data for this survey originated from the baseline of the R-SWITCH Ethiopia study, a cluster-randomized controlled trial (cRCT) evaluating the impact of an integrated intervention package. The R-SWITCH intervention was designed to enhance the detection and treatment coverage of severe acute malnutrition (SAM) across the full continuum of care prevention, screening, referral, treatment, and relapse prevention at household, community, and facility levels in two Ethiopian woredas. The package included monthly group behavior change communication sessions led by Alliance for Development (AFD) groups on IYCF, health, and WaSH, with food-based recipe demonstrations; family-led weekly MUAC screening by trained caregivers plus active and passive screening at health services; and expanded admission criteria (MUAC < 115 mm, edema, or WAZ < −3). The intervention was delivered through health posts, AFD groups, and community leaders within a cluster RCT design.

The survey enrolled 686 children aged 6–59 months identified with SAM or enrolled in SAM treatment across 40 health post catchment areas in two distinct livelihood zones: (i) the agrarian Kersa woreda, Jimma Zone, Oromia Region, and (ii) the agrarian and pastoral Jeldesa woreda, Dire Dawa City Administration. Sites were purposefully selected based on high SAM burden, accessibility, and programmatic diversity using data from 2015–2021. Kersa, a rural coffee-producing woreda located about 357 km southwest of Addis Ababa, serves approximately 40,567 children under five, while Jeldesa (about 3,956 under-fives) represents typical agrarian and pastoral livelihood systems. Of 44 health posts/kebeles, 40 eligible units (with at least two HEWs) across both woredas participated in the survey. Woredas are key administrative districts, while kebeles are the smallest, community-level administrative units responsible for basic services, security, and development.

Enumerator teams performed door-to-door anthropometric screening of all children aged 6-59 months. SAM eligibility was determined by mid-upper arm circumference (MUAC) <115 mm, weight-for-length Z-score (WLZ) < -3 (2006 WHO standards), bilateral pitting edema, or existing enrollment in OTP. Children with congenital anomalies preventing accurate measurement or caregiver non-consent were excluded.

### Sample size

The sample size was calculated for the R-SWITCH Ethiopia baseline survey a cluster-randomized controlled trial (cRCT) based on projected SAM OTP treatment coverage among children aged 6–59 months. Due to limited recent data, conservative baseline coverage of 50% was assumed. Detecting a 15% absolute increase (50% to 65%) required 40 clusters (health posts) with 90% power, 5% significance level (α = 0.05), design effect of 1.5, cluster size of 30 children, and 10% non-response adjustment, yielding a target of 686 children with SAM or enrolled in OTPs. Within this cRCT framework, all eligible children aged 6–59 months across clusters underwent assessment of nutritional status, caregiver knowledge, IYCF practices, and WaSH practices. During the May1–September30 2024 baseline survey in Jeldesa and Kersa woredas, EPHA and IFPRI teams utilized Survey Solutions on Android tablets for face-to-face data collection. The dataset used in this study was accessed for research purposes on November 10, 2024, under a formal data-sharing agreement with the International Food Policy Research Institute (IFPRI) and the Ethiopian Public Health Association (EPHA). The data provided to the authors were fully anonymized and did not include any personally identifiable information; therefore, individual participants could not be identified during or after data collection. Trained enumerators gathered two primary datasets: (1) anthropometric screening of approximately 28,000 children aged 6–59 months; and (2) detailed household, caregiver, and child-level information from 686 children identified with SAM.

### Data collection tools, procedures and measurements

Caregiver knowledge (primary outcome) was assessed using a standardized 32-item questionnaire spanning five domains: IYCF (breastfeeding/complementary feeding), child health, CMAM, and SAM screening. Composite scores were calculated for each domain and combined into an overall caregiver knowledge score. The secondary outcome, IYCF indicators followed WHO/UNICEF 2021 standards (21): (1) continued breastfeeding, (2) timely complementary food introduction, (3) minimum dietary diversity (MDD ≥5 out of 8 food groups), (4) minimum meal frequency (MMF) by age, (5) minimum acceptable diet (MAD: meeting both diversity and frequency), (6) egg/flesh foods, (7) sweet beverages, (8) sentinel unhealthy foods, and (9) zero vegetable/fruit (ZVF). All indicators used dichotomous scoring (0/1) based on 24-hour recall. Each indicator was scored dichotomously (0/1) outcome based on 24-hour recall. All measures adhered to WHO 2021 definitions(22). Water, Sanitation, and Hygiene (WaSH) variables assessed treated/improved drinking water, availability of hand washing station with soap, and primary water source (improved vs unimproved). Determinant variables included maternal factors (age, education, literacy, maternal knowledge, etc). Maternal depression was screened using the Edinburgh Postnatal Depression Scale (EPDS), a validated 10-item questionnaire for perinatal depression symptoms(23). For analysis, EPDS scores were dichotomized using established cut-offs: possible depression (EPDS ≥ 10) versus no/less likely depression (EPDS < 10)(23). Caregiver community involvement was assessed using a 10-item scale measuring participation in women’s discussion groups (health, nutrition, school problems, community issues), where higher scores indicated greater involvement. Caregiver mobility was assessed using a 10-item scale measuring permission requirements for daily activities (visiting family, markets, health services, community issues), where higher scores indicated greater household restrictions. Caregiver decision making autonomy (independent decisions (made alone, without spousal/others approval) reported by caregivers across key domains: child feeding practices, healthcare seeking, food/livestock purchasing, and household expenditures. This was scored on a 13 scale (higher scores indicating greater autonomy), following validation against standard DHS Women’s Empowerment indices), marital status (married/living with husband), income-generating activities, Caregiver stigma perceptions were measured using a 9-item questionnaire assessing fears about seeking care at health centers for malnourished children (e.g., "people will think the family has no food," "cannot afford care," "or is ‘bad’ parents," "household is destitute/dirty," "will be looked at badly," "health worker will blame us," and "health worker will talk down to us"). The analysis examined these perceptions among caregivers of children with SAM, focusing on reported fear or shame about visiting HEWs and stigma related to AFD/HEW services. Child characteristics (age, sex, anthropometry), household wealth, and household (HH) food insecurity assessed using the validated Household Food Insecurity Access Scale (HFIAS) were also evaluated(24).

Anthropometric measurements were standardized via three training sessions featuring repeated assessments on 10 children, ensuring enumerator teams achieved data accuracy and reliability. The survey collected IYCF indicators, food insecurity, wealth index, hygiene practices, child health status, and livelihood information. All instruments were translated into Amharic and Oromifa, back-translated for validation, pretested, and deployed following 14 days of standardized enumerator training.

### Data Management and Quality Control

Field coordinators oversaw trained enumerators who used WHO-standard guidelines to gather anthropometric data(25). Data collectors received standardisation training and were instructed to take all necessary safety measures, such as calibrating equipment, setting it correctly, reading values accurately, and accurately entering data. Every day, the data was reviewed for completeness, and any flaws that were found were fixed before it was safely and anonymously uploaded to a password-protected server. Before the main survey started, a pilot research was carried out in Manna Woreda using an informed questionnaire and procedure modifications.

### Statistical Data Analysis

Maternal knowledge of IYCF, child health, SAM screening, and CMAM was assessed using R-SWITCH baseline survey data from 40 clusters (health posts) across rural agrarian and pastoralist settings in Ethiopia. Standardized questionnaires were adapted from WHO IYCF indicators, EDHS modules, and UNICEF multiple indicator cluster surveys. Instrument validity was established through content validity by five independent experts and pilot testing (n=50; 7% of target sample) in adjacent districts(26). The questionnaire was developed in English, forward-translated into Oromiffa and Amharic, and back-translated by certified linguists to ensure linguistic equivalence. Internal consistency reliability was confirmed with Cronbach’s α = 0.80 across domains. Household, caregiver, and child characteristics were summarized by woreda and livelihood zone using descriptive statistics (Stata v17). Composite knowledge scores across five domains were computed and dichotomized at the mean (≥mean vs. <mean) by woreda.

Two complementary analyses were conducted among caregivers of children aged 6–59 months newly identified with SAM or enrolled in OTPs. First, determinants of caregiver knowledge (dependent variable) were identified using bivariate screening (P<0.25*P*<0.25) and multivariable mixed-effects linear regression (P<0.05*P*<0.05 for retention; coefficients reported as change in knowledge score). Second, associations between knowledge (independent variable) and IYCF/WaSH practices (dependent variables) were examined using mixed-effects Poisson regression models with robust standard errors to estimate incidence rate ratios (IRRs) for each practice (e.g., continued breastfeeding, minimum dietary diversity, improved water source, handwashing). These associations were illustrated as forest plots.

Eight binary IYCF indicators were analyzed for children aged 6–23 months, as these are tied to the complementary feeding period and have age-specific meanings (27). Dietary diversity served as the main diet quality indicator for children aged 24–59 months (28). WaSH practices were assessed for all children aged 6–59 months. Clustering was addressed using kebele as random effects and woreda as fixed effects.

### Ethical Considerations

In accordance with national research ethics guidelines, the Ethiopian Midwives Association IRB granted ethical approval (Protocol Ref: EMwA-IRB-SOP-/010/4-24). Anthropometric measurements followed WHO protocols with ethical safeguards: trained professionals took gentle measurements with parental consent; no child was forced. All screened SAM cases were immediately referred to nearest CMAM OTP per national guidelines. To guarantee openness and community support, the goal of the study was explained to local health officials and community leaders prior to fieldwork. Before conducting in-depth interviews, all caregivers provided written informed consent; oral consent was obtained for quick anthropometric screening. The research protocol can be found at (https://ichgcp.net/clinical-trials-registry/NCT06380504).

## Results

### Socio-demographic characteristics

All caregivers who participated provided written informed consent for survey interviews, with zero refusals. Among 686 caregivers in agrarian (607) and pastoralist (79) settings, 96.1% were biological caregivers, average age 28.8 years (± 7.0 SD). years; 93.0% were married. Only 27.9% had formal education, higher in pastoralist (75.9%) than agrarian (43.9%) areas. Children had an average age of 19 months (±13.0SD) (Table1).

**Table 1:**
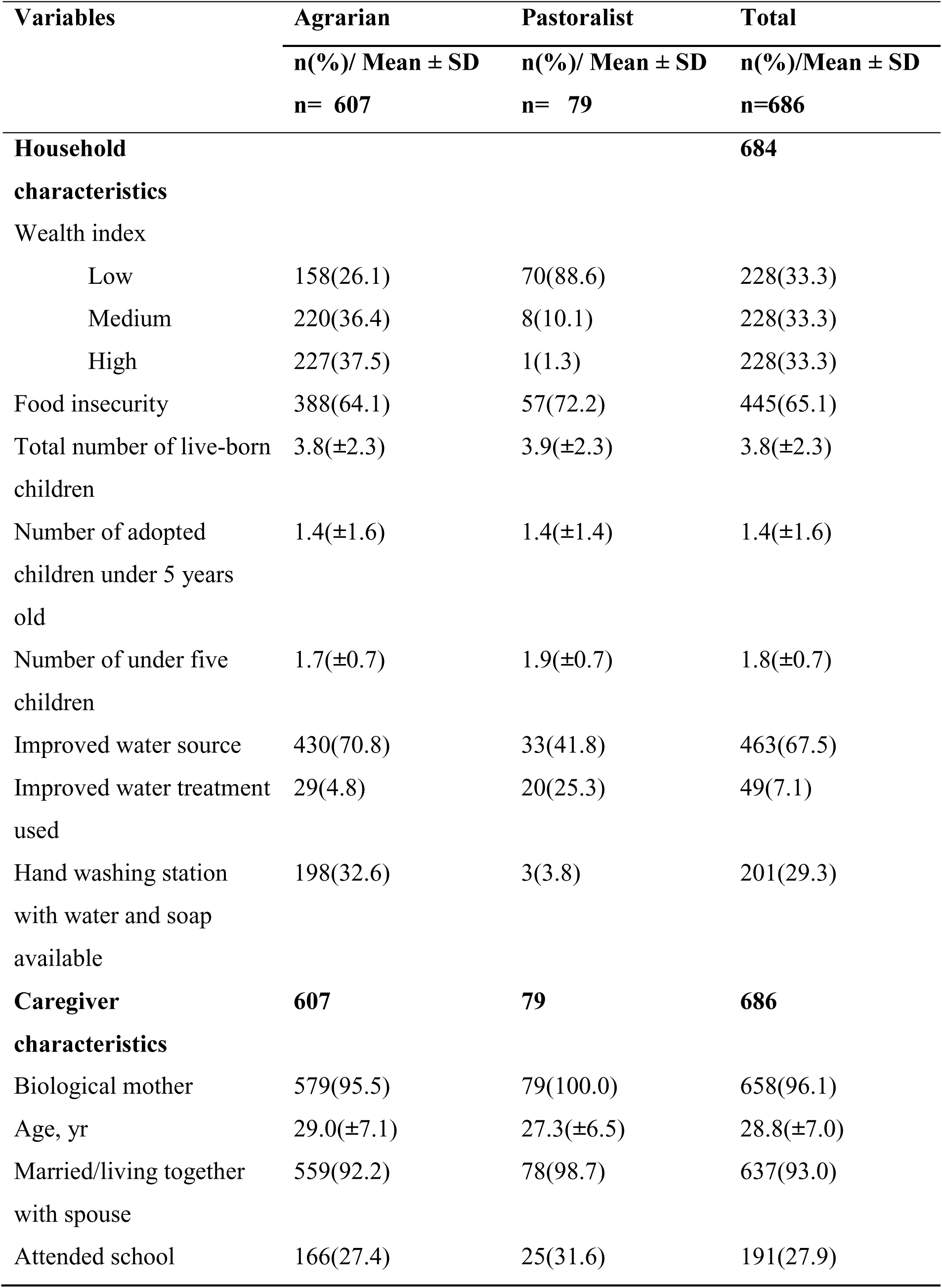
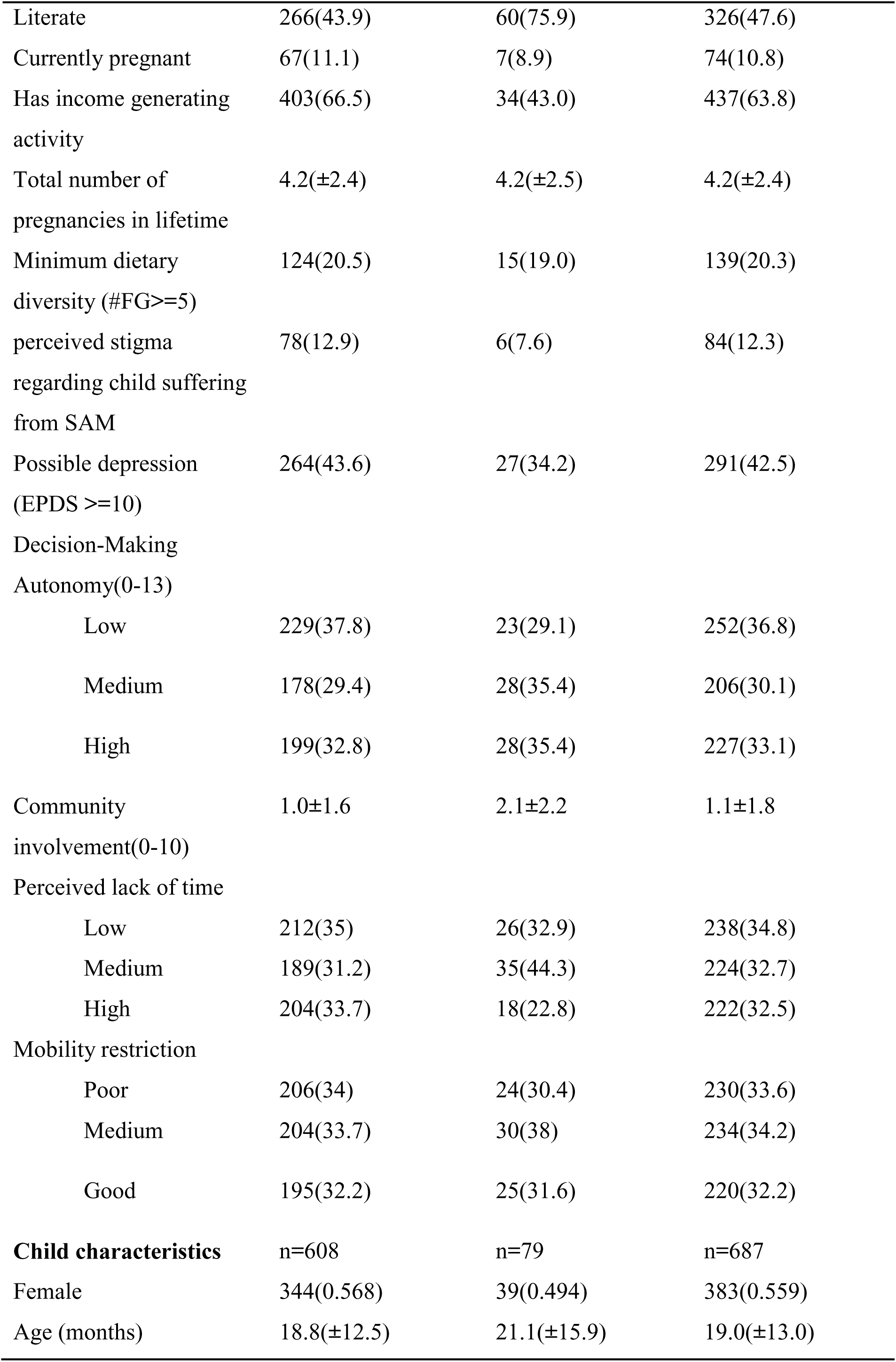
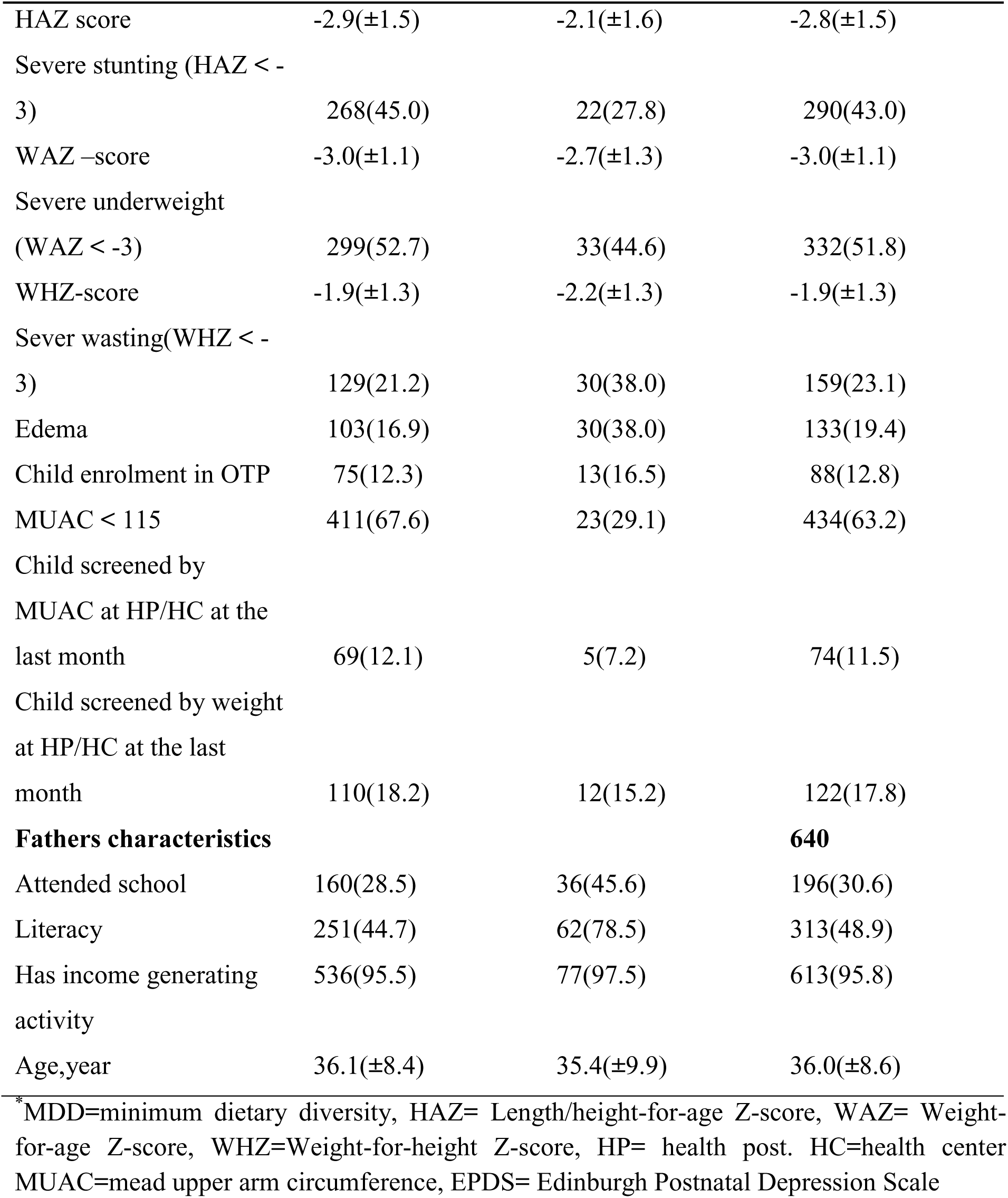
Characteristics of Household, Caregiver, Children and father in Agrarian and Pastoralist Settings.

### Caregiver Knowledge of IYCF practices, Child Health, CMAM, and SAM screening

Out of 686 caregivers assessed, 44.5% scored at or above the mean knowledge score (9.5 ± 3.9/32), with pastoralist mothers scoring slightly higher than agrarian mothers. Breastfeeding knowledge was moderate (66.6%), but complementary feeding knowledge was low (25.3%).

**Table 2:**
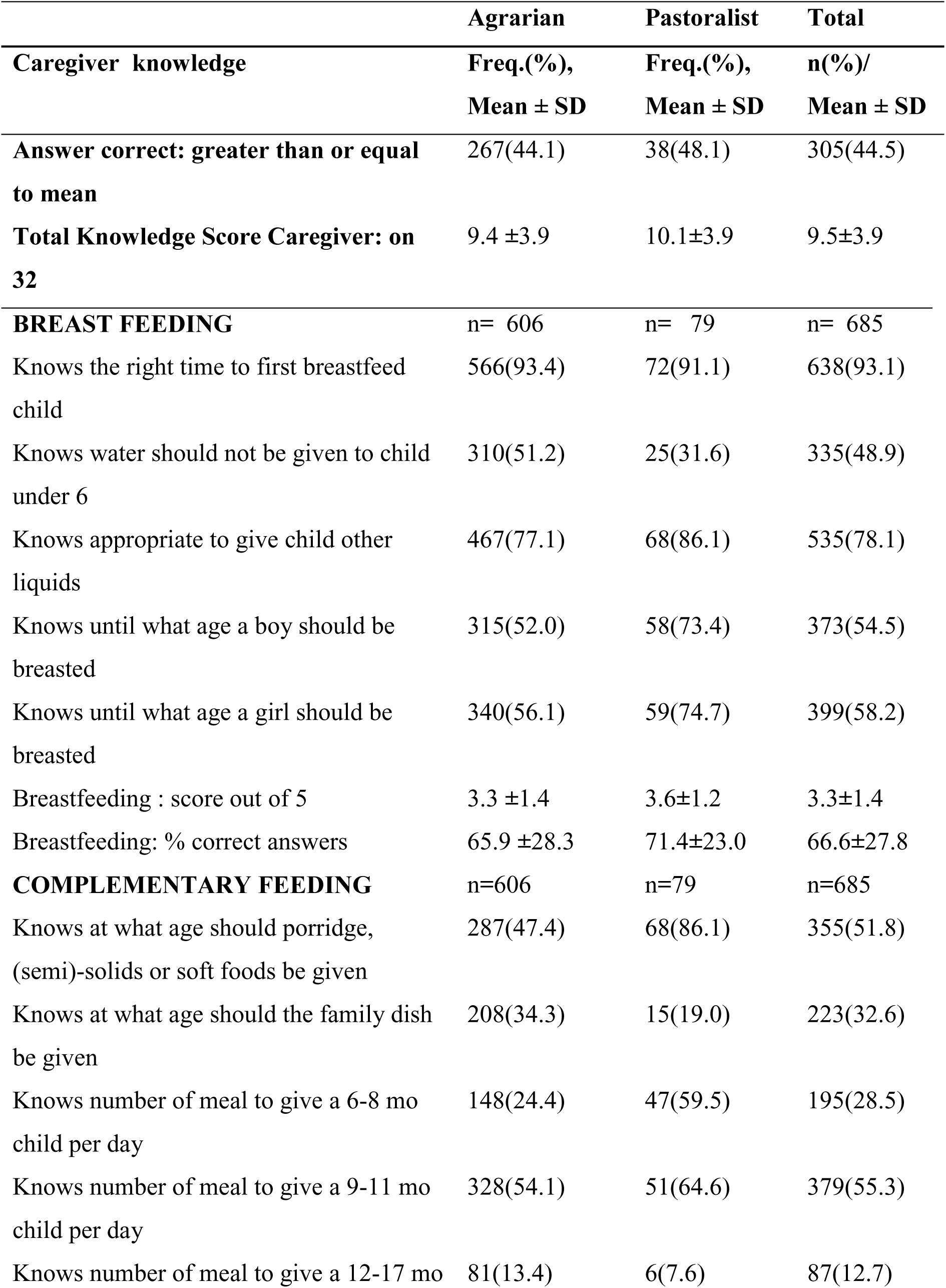
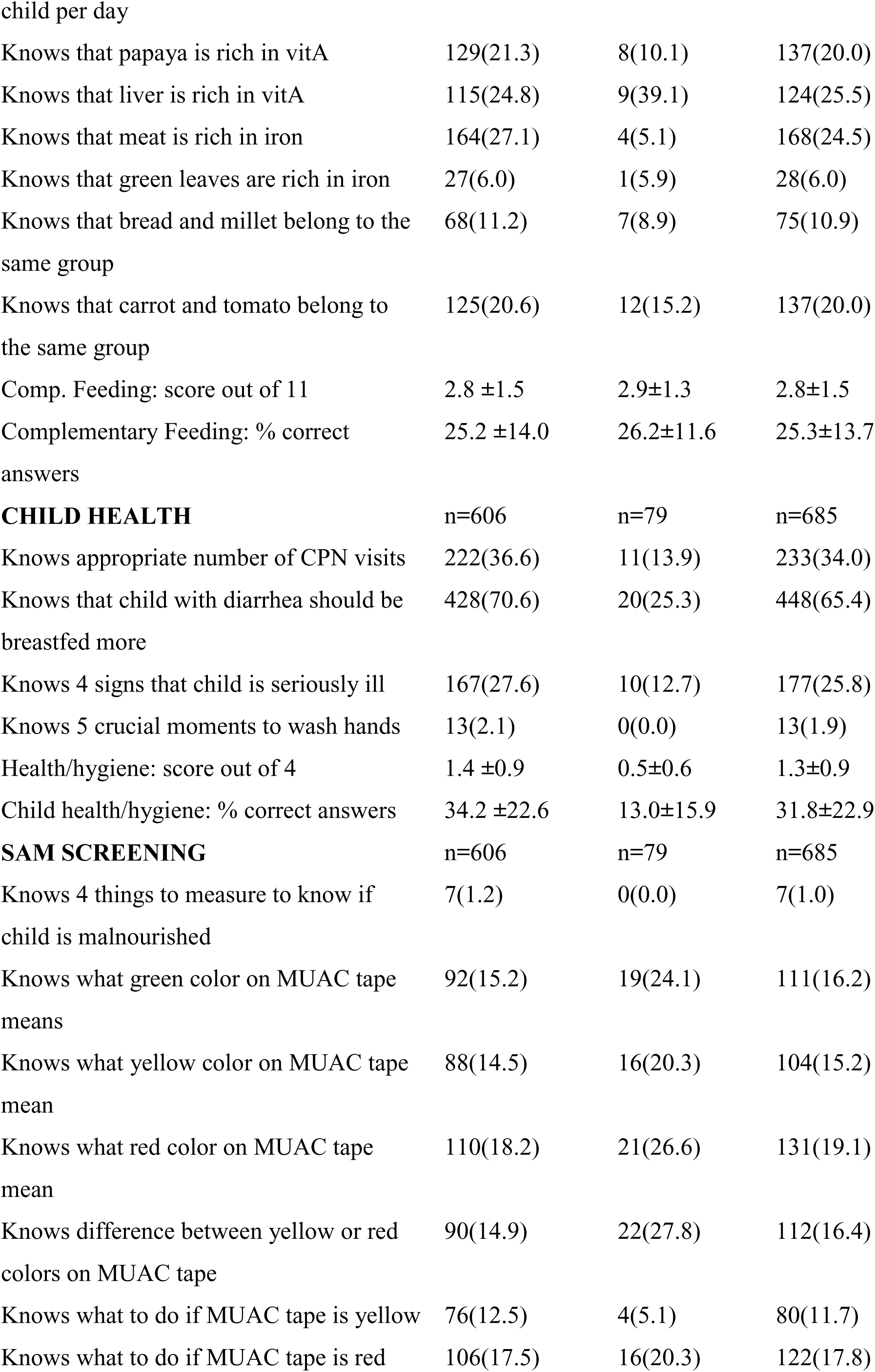
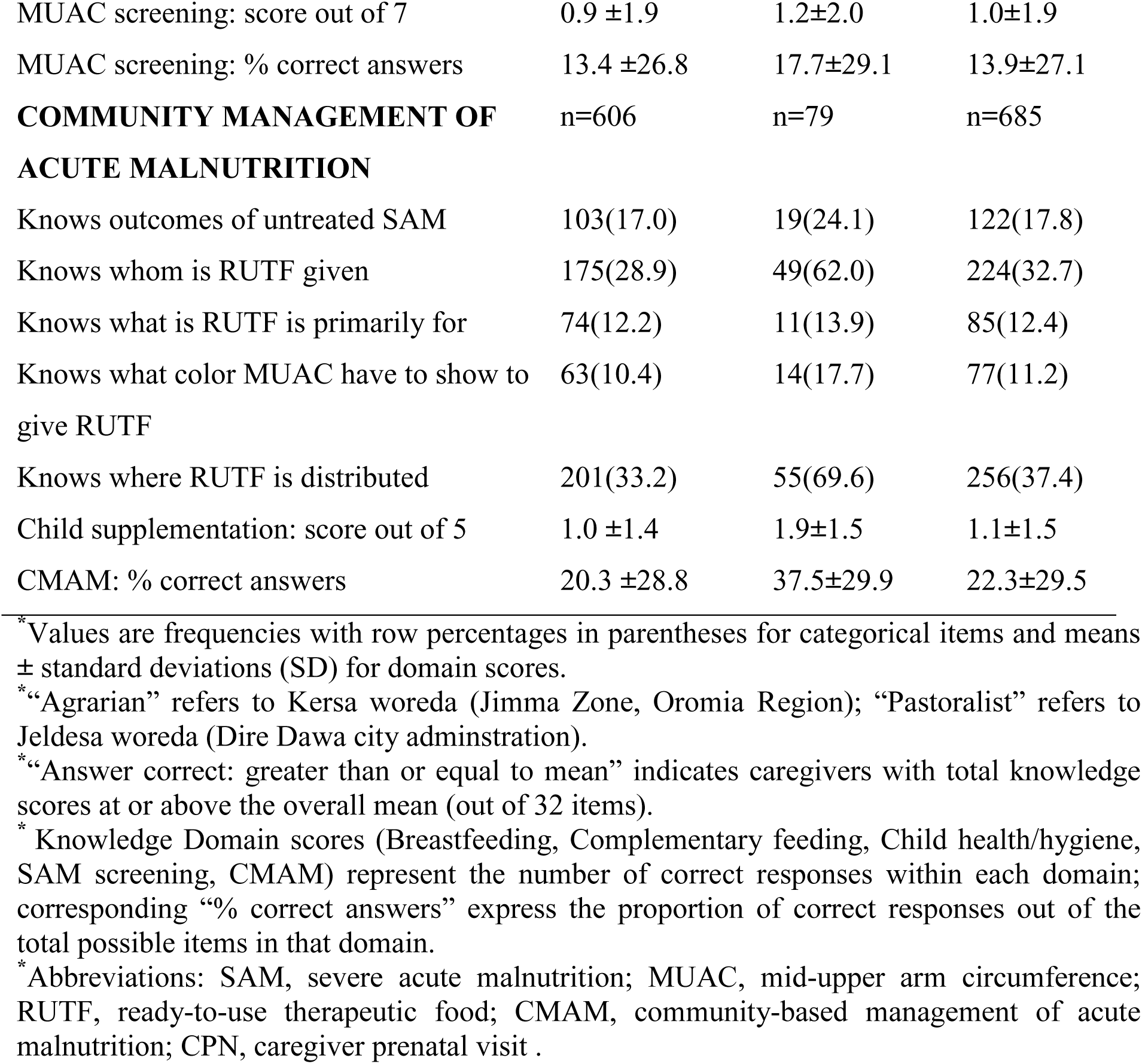
Domains of Caregiver Knowledge among Children Aged 6–59 months identified with SAM and admitted to SAM OTP in Agrarian and pastoral Ethiopia.

### Association between Caregiver Knowledge and IYCF Practices

Caregiver knowledge was positively associated with several IYCF practices among children aged 6–59 months with SAM (Fig. 1). A forest plot was used to illustrate the association between caregiver knowledge and IYCF practices. Each one-unit increase in knowledge score was associated with higher likelihood of achieving minimum dietary diversity (MDD, IRR=1.50, 6–23 mo) and minimum acceptable diet (MAD, IRR=1.63, 6–23 mo), plus greater dietary diversity among older children (24–59 mo, IRR=1.70). Higher caregiver knowledge was also associated with a lower likelihood of children having zero vegetable or fruit intake (IRR = 0.77).

**Figure 1:**
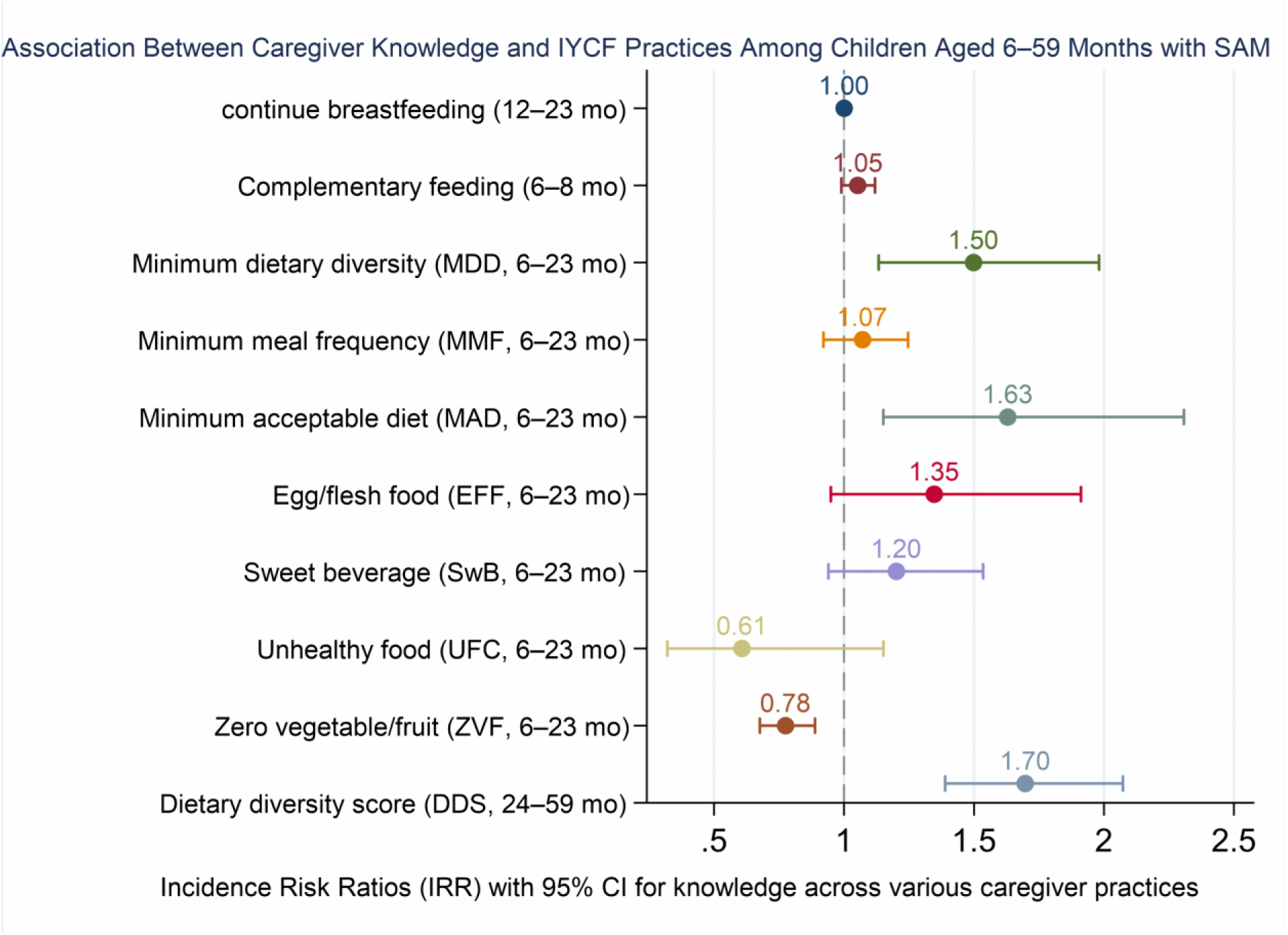
The association between the total knowledge scores of caregivers with the IYCF practices in their children identified with SAM and admitted to SAM OTP. * Incidence rate ratios (IRRs) and 95% confidence intervals for the caregiver knowledge score, estimated from mixed-effects Poisson regression models adjusted for clustering, show its effect on each infant and young child feeding (IYCF) outcome among children 6–59 months with SAM. Associations are statistically significant when the confidence interval excludes IRR=1.0.

### Association between caregiver knowledge and WaSH practices

Caregiver knowledge showed positive associations with key WaSH practices among children aged 6–59 months with SAM (Fig. 2). A forest plot was used to illustrate the association between caregiver knowledge and WaSH practices. Each one-unit increase in knowledge score associated with greater likelihood of using improved water sources (IRR=1.19), improved water treatment (IRR=2.02), and having a handwashing station with soap available (IRR=1.41).

**Figure 2:**
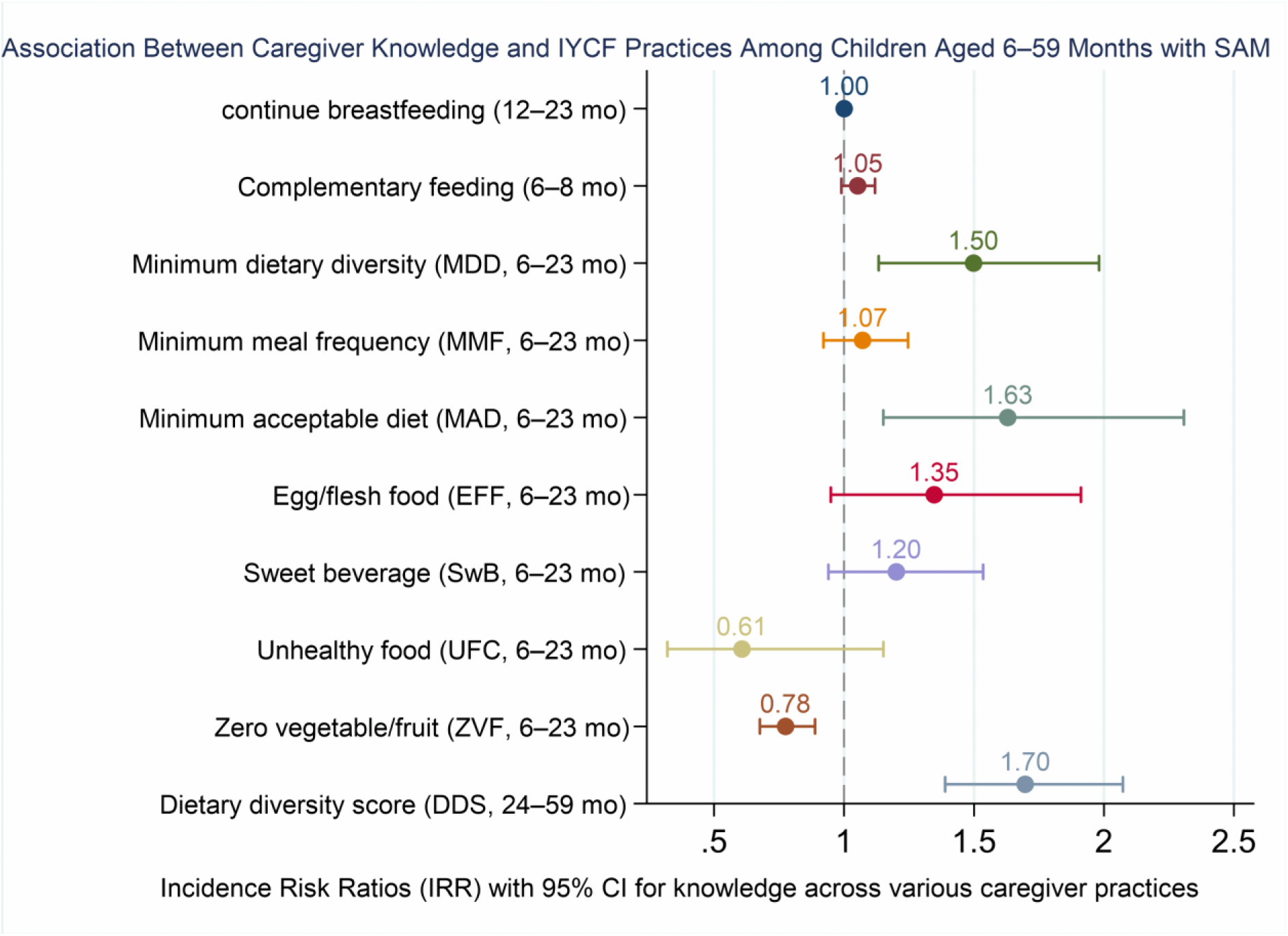
The association between the total knowledge scores of caregivers with the WaSH practices in their children identified with SAM and admitted to SAM OTP. * Incidence rate ratios (IRRs) and 95% confidence intervals for the caregiver knowledge score, estimated from mixed-effects Poisson regression models adjusted for clustering, show its effect on each WaSH practices among children 6–59 months with SAM. Associations are statistically significant when the confidence interval excludes IRR=1.0.

### Determinants of knowledge about IYCF practices, child health, CMAM, and SAM screening among caregivers of children aged 6–59 months

The multivariable analysis showed that caregiver literacy was associated with higher caregiver knowledge scores (β = 3.8, 95% CI: 0.7–7.0; p= 0.016). Current pregnancy was also associated with higher knowledge scores (β = 3.8, 95% CI: 0.7–7.0; p= 0.018). A higher household wealth index was strongly positively associated with increased knowledge scores (β = 11.8, 95% CI: 3.6–20.1; p=0.005), and having the child weighed at a health facility in the past month was likewise associated with higher knowledge (β = 8.9, 95% CI: 3.5–14.2; p≤0.001). In contrast, possible depression was associated with lower caregiver knowledge scores (β = −0.3, 95% CI: −0.5–0; p= 0.015) (Table 3).

**Table 3:**
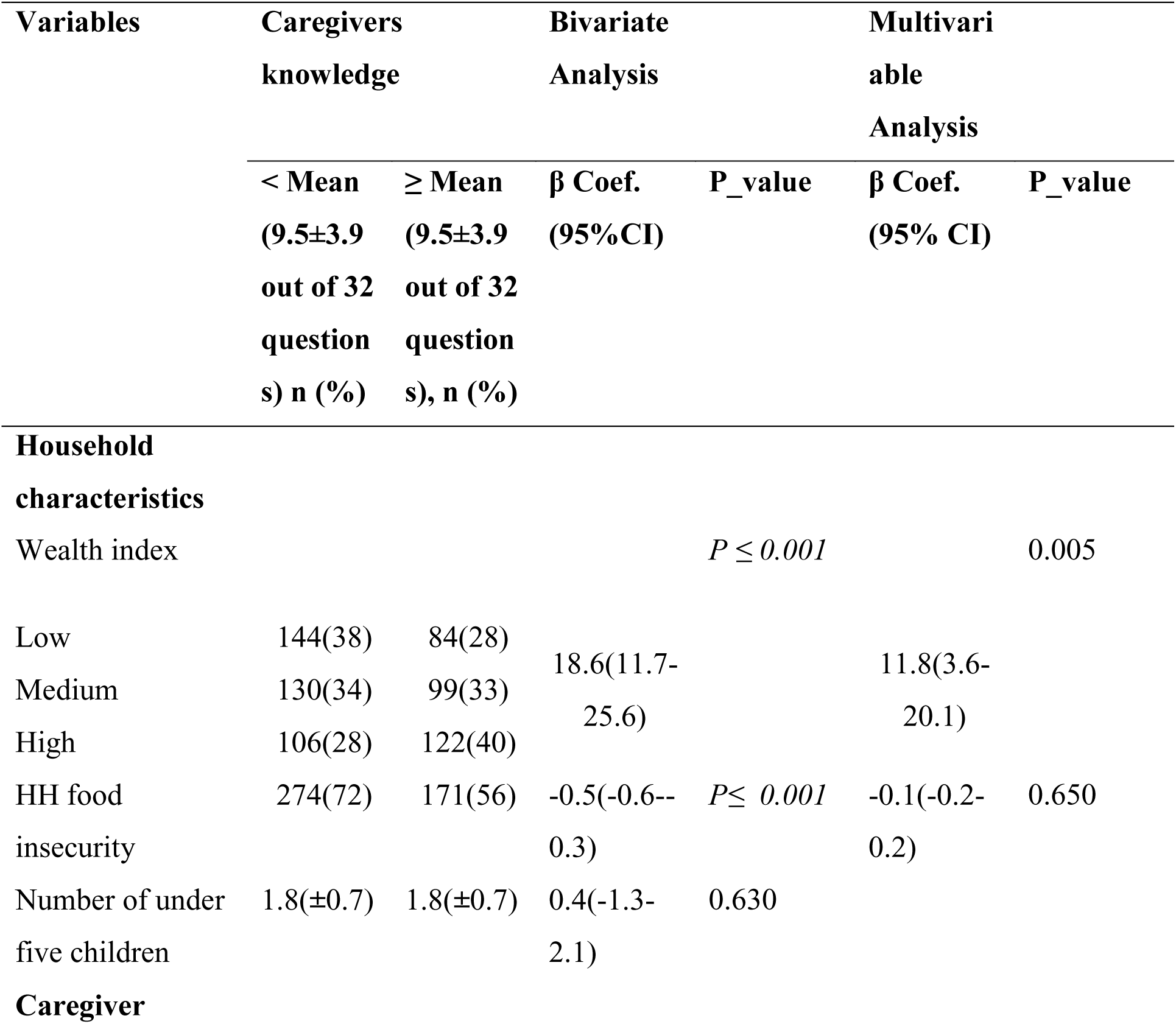
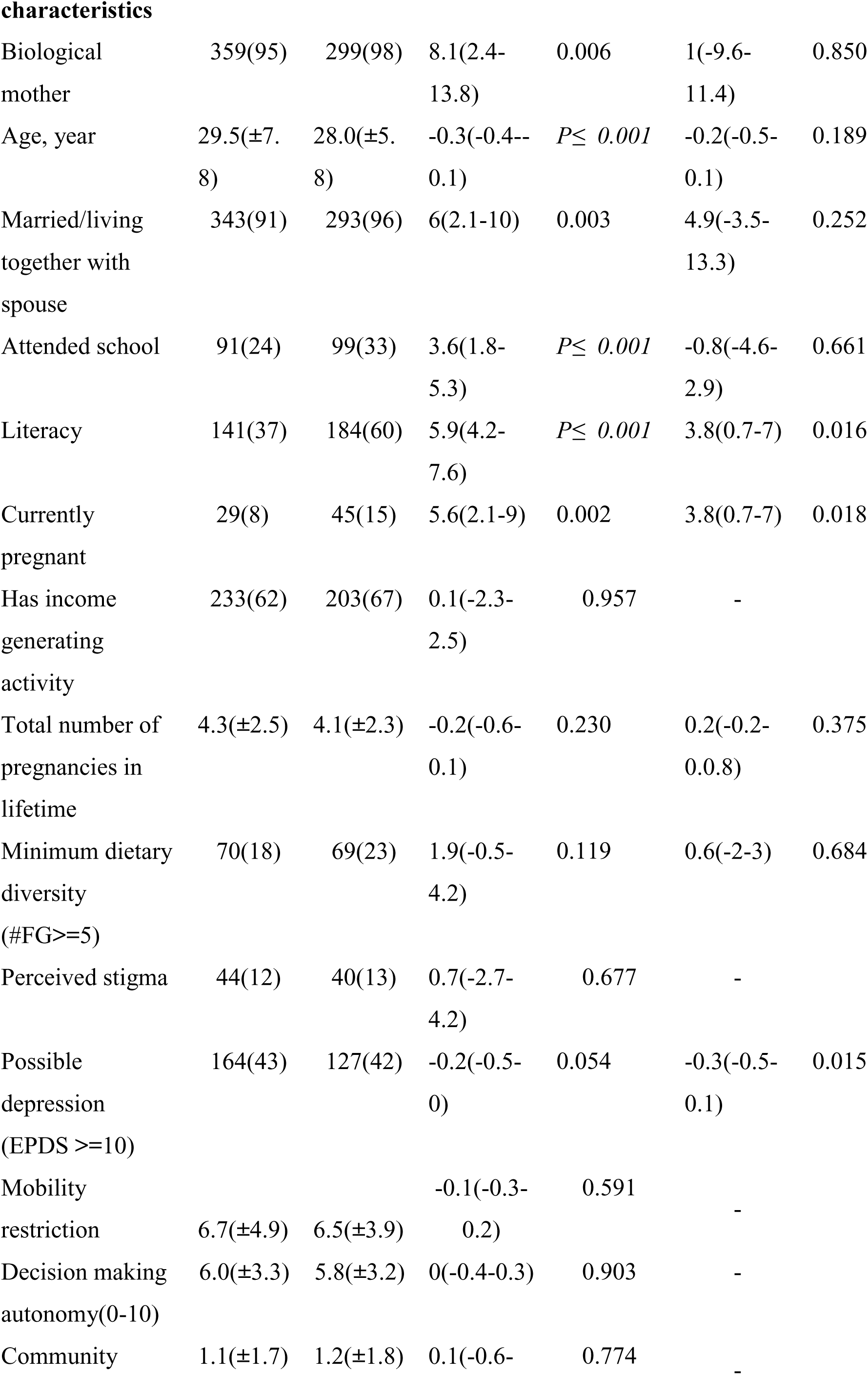
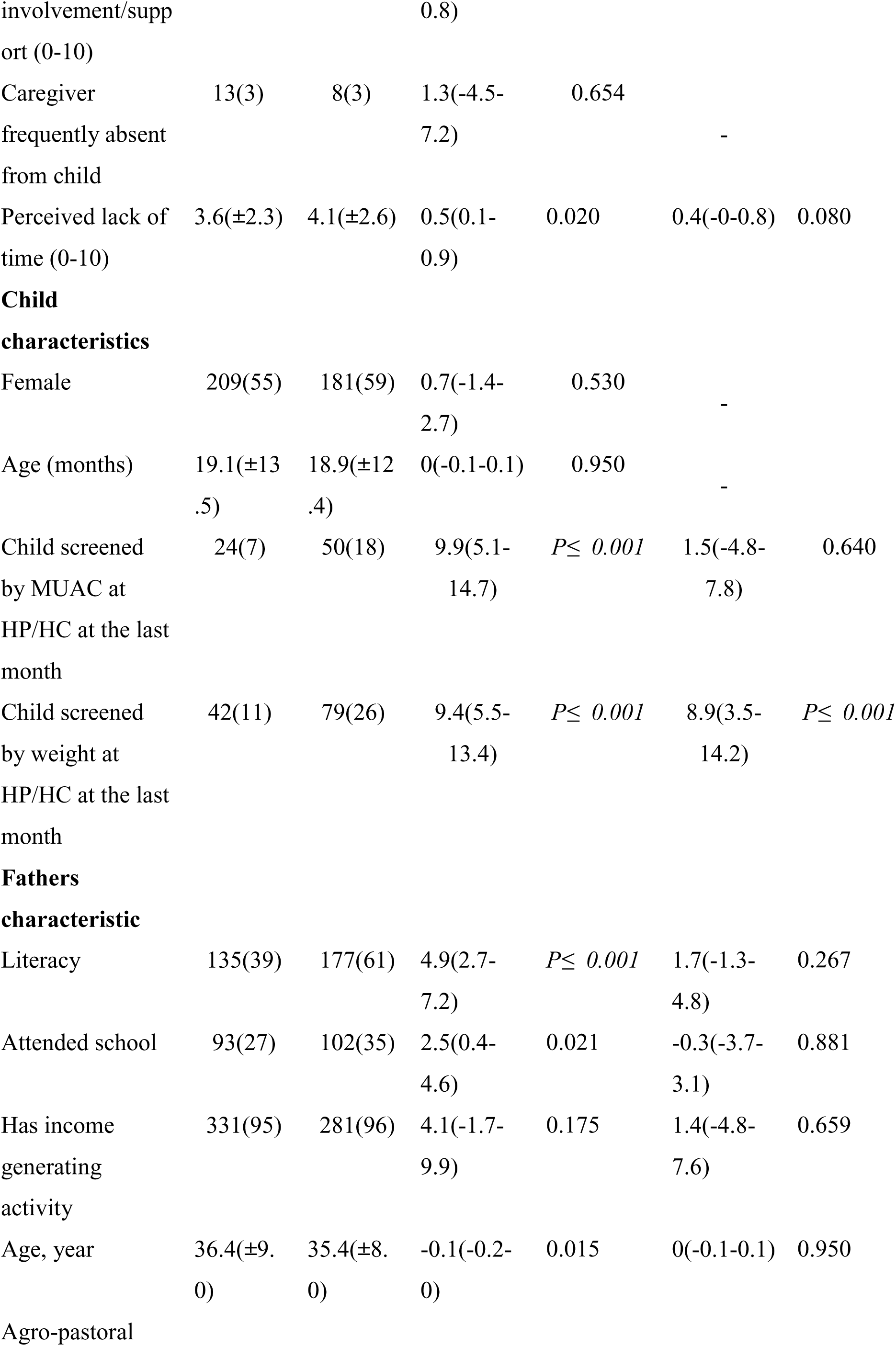
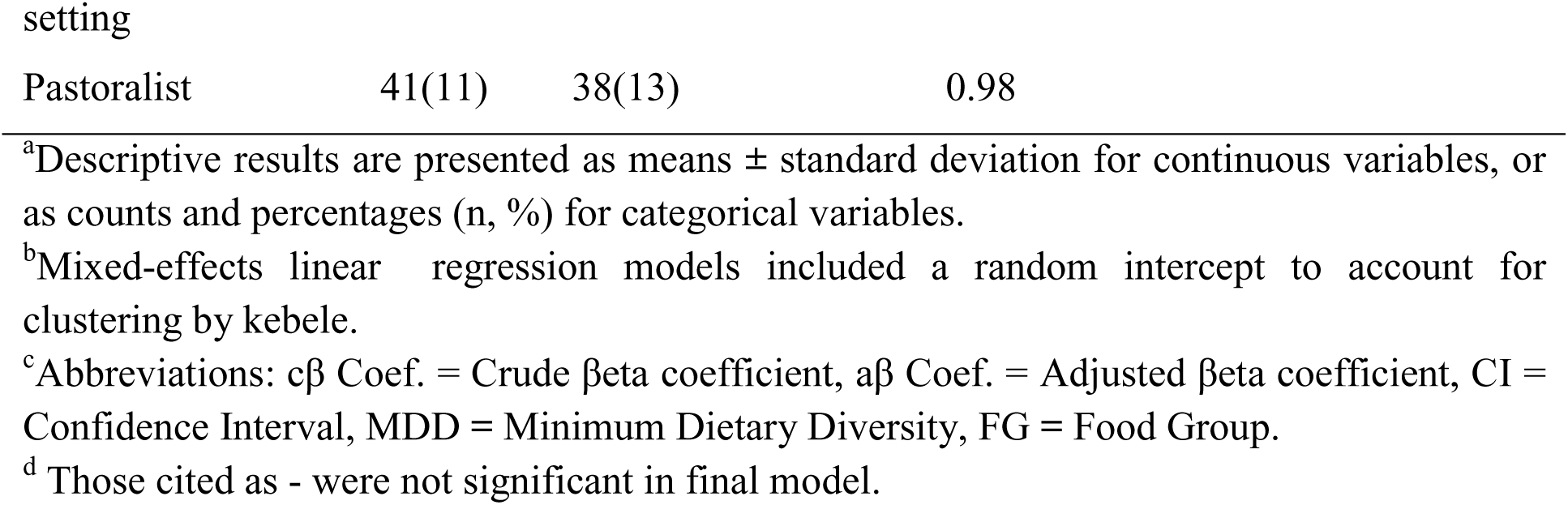
Multivariable analysis of determinants of caregivers’ knowledge about IYCF practices, child health, CMAM, and SAM screening among caregivers of children aged 6–59 months in agrarian and pastoral settings, Ethiopia, 2024.

## Discussion

This study primarily examined caregiver knowledge across five domains breastfeeding, complementary feeding, child health, SAM screening, and CMAM among children aged 6– 59 months with SAM in agrarian and pastoralist settings, along with its determinants. Secondarily, it assessed associations between this knowledge (specifically breastfeeding, complementary feeding, child health, SAM screening, and CMAM) and IYCF/WaSH practices. Among 686 caregivers from agrarian (Kersa) and pastoralist (Jeldessa) areas, only 44.5% achieved scores at or above the mean total knowledge score (9.5 ± 3.9 out of 32), with slightly higher levels in pastoralist (48.1%) versus agrarian (44.1%) settings.

### Association between Caregiver Knowledge and IYCF/WaSH Practices

This study found a significant positive association between caregivers’ knowledge and MDD achievement among children aged 6–23 months with SAM. Hypothesized pathways include: complementary feeding knowledge facilitating age-appropriate nutrient-rich food inclusion; breastfeeding knowledge supporting continued breastfeeding alongside solids; and SAM screening awareness prompting proactive dietary diversification to prevent deterioration. These findings align with evidence from Ethiopia, Sub-Saharan Africa, and South Asia showing caregiver nutrition knowledge consistently predicts better child dietary diversity and feeding practices (29–32).

This study revealed a significant association between caregiver knowledge and MAD achievement among children aged 6–23 months with SAM, positioning caregivers as key mediators translating IYCF principles proper meal frequency and dietary diversity into practice for nutritional recovery. Better-informed caregivers, particularly those counseled at health posts with good literacy, breastfeeding knowledge, complementary feeding timing, and infant feeding frequency understanding, likely drive MAD success. These findings align with Ethiopian evidence where complementary feeding and infant feeding practice knowledge independently predict MAD(33,34).

Caregiver knowledge was inversely associated with zero fruit/vegetable consumption of children aged 6-23month with SAM, consistent with evidence linking maternal education, ANC counseling, wealth index and responsive feeding practices to reduced zero-intake prevalence in East Africa(35,36).

This study found caregivers’ knowledge strongly associated with improved water sources, household water treatment, and hand washing stations with soap among children aged 6–59 months with SAM which may be attributed to Ethiopia’s Health Extension Program’s integrated approach through frequent health post visits and HEW communication. Caregivers receiving simultaneous nutrition/WaSH counseling during SAM screening/CMAM recognize the nutrition-WaSH nexus (diarrhea, nutrient loss, malnutrition-infection cycle), prioritizing dietary diversity alongside water safety. This health literacy positions caregivers as proactive agents linking feeding and hygiene practices. Supporting evidence from households with higher awareness in Southern Ethiopia indicates that these households were more likely to treat drinking water, underscoring the association between knowledge and WaSH practices (37). WaSH education from Health Extension Workers similarly improves water treatment, latrine use, and handwashing facilities in Jimma Zone(Alemu et al., 2023; Soboksa et al., 2021).

### Determinants of knowledge about IYCF practices, child health, CMAM, and SAM screening among caregivers of children aged 6–59 months

Higher caregiver literacy was strongly associated with better knowledge scores, which may be due to greater household wealth enabling education access, frequent health post/health center visits for SAM screening/child follow-up, and exposure to diverse information sources. Enhanced literacy facilitates comprehension of medical guidance, promoting timely preventive actions and effective service utilization. These findings align with evidence that parental health literacy predicts health knowledge outcomes (38) and are consistent with Ethiopian (39), confirming literacy as a key determinant of health knowledge and care service utilization.(40).

This study found that pregnant or multigravida caregivers of children with SAM demonstrated greater knowledge than non-pregnant caregivers. This may stem from the interplay of education, counseling, prior caregiving experience, and frequent contact with health extension workers during pregnancy, which provides enhanced access to information. These results align with Ethiopian studies showing that caregivers with more pregnancies tend to be more knowledgeable (41) and that multiparous caregivers possess greater expertise than primiparous ones (39), ANC visits provide pregnant women with frequent nutrition counselling, enhancing infant feeding knowledge and offering a key chance to boost caregiver education.

The household wealth index was associated with caregiver knowledge; caregivers from wealthier households showed higher nutrition and IYCF knowledge. This may be explained by their greater access to transportation for frequent health post/center visits, media exposure, quality education, timely growth monitoring and promotion (GMP), opportunities to practice their knowledge, and improved communication with health extension workers. This is consistent with evidence indicating that better economic status is associated with greater access to information and health services(42,43). Further support comes from studies in Pakistan, which also link caregivers’ socioeconomic standing to their levels of nutritional awareness(44).

Children having been weighed at a health post or health center in the preceding month were strongly associated with increased caregiver knowledge, indicating that contact with GMP services was linked to higher levels of caregiver knowledge. Regular weighing is typically accompanied by counseling on child feeding, growth curves, and signs of undernutrition, which were associated with greater caregiver understanding of nutrition and health. Studies from Ethiopia and Ghana report that exposure to GMP sessions and primary health care was positively associated with higher maternal knowledge scores on child feeding and growth, as well as with better reported practices(45,46).

This study found that potential depression was linked to a slight but significant decrease in caregivers’ knowledge, even though no quantitative Ethiopian studies have yet connected caregivers’ knowledge to psychological well-being variables such as stress, anxiety, and depression. Similar studies document substantial mental health burdens among caregivers of malnourished children in Kenya, which reduced their awareness capacity (47). This finding is also supported by a Chinese study showing that stress and depression is inversely related to caregiver knowledge, as it promotes better coping and a sense of fulfillment that enhances caregiving quality (48).

This study had several strengths and limitations. The data were drawn from caregivers of children aged 6–59 months with SAM or enrolled in SAM treatment a population-representative sample from two distinct settings, a notoriously difficult group to capture given their low prevalence, which required screening approximately 28,000 children in this study. The sample spanned two distinct woredas, though the pastoralist woreda contributed a notably smaller sample size due to its smaller overall population. Data were collected through house-to-house surveys that captured detailed child, caregiver, and household characteristics, including direct observation of assets, wealth indicators, and WaSH facilities; caregiver knowledge was comprehensively assessed across IYCF, child health, SAM screening, and CMAM domains. However, several limitations should be noted. The cross-sectional study design precludes causal inference. Self-reported data may be subject to social desirability bias. The nomadic lifestyle of pastoralist households also limited opportunities for repeated visits, resulting in missing paternal data.

## Conclusion

Caregiver knowledge across IYCF, child health, SAM screening, CMAM, and WaSH domains was suboptimal in these high-burden agrarian and pastoralist settings, yet consistently associated with better feeding and hygiene practices among children with or recovering from SAM. Higher knowledge linked to improved dietary quality (MDD, MAD, and reduced zero vegetable/fruit) and safer WaSH behaviors (treated/improved water, functional handwashing stations). Key positive determinants included literacy, pregnancy/multigravida, higher household wealth, and child weighing at a health post/center; depressive symptoms emerged as a significant negative correlate. These findings highlight the need to integrate structured, context-specific caregiver education and psychosocial support into CMAM, GMP, and primary health services these settings communities to bolster IYCF, WaSH practices and mental health support, ultimately reducing SAM and under nutrition burdens in Ethiopia.

## Data Availability

The minimal data set supporting the findings of this study is available upon reasonable request from the International Food Policy Research Institute (IFPRI) and the Ethiopian Public Health Association (EPHA), which securely manage the data on password?protected servers. Data cannot be shared publicly because access is restricted to authorized researchers who meet the criteria for use of confidential data. Requests should be directed to, and and will be considered by the relevant data?access committee in accordance with ethical and institutional requirements.

## Acknowledgements

The authors express their profound gratitude to Haramaya University, Jimma University, the United Nations International Children’s Emergency Fund (UNICEF), the Ethiopian Public Health Association (EPHA), and the International Food Policy Research Institute (IFPRI) for their important assistance and cooperation. We would especially like to thank the Melinda Gates Foundation for their generous funding and dedication to furthering the field of maternal and child nutrition research. These institutions’ collaboration and resources have been essential to this study’s successful conclusion.

## Author contributions and consent for publication

Each author contributed to the conception of TB, LH, MA, and AH. The research project was designed by TB, LH, MA, and AH. In formal analysis and interpretation, MA, LH, MT, TB, and TF. In supervision, TB, LH, MA, BB, DT, and AH. The original paper was written by MA. Writing: Editing and review. LH, DT, MT, TB, and TF The final text was reviewed and approved by all authors. MA is in charge of writing the article and submitting it.

## Availability of data and materials

Only authorised researchers have access to the password-protected servers run by the International Food Policy Research Institute (IFPRI) and the Ethiopian Public Health Association (EPHA), which securely handle and preserve the data.

